# Global Warming and Neurological Practice: Systematic Review

**DOI:** 10.1101/2020.12.18.20248515

**Authors:** C Peinkhofer, M Amiri, MH Othman, T De Vecchi, V Nersesjan, D Kondziella

## Abstract

**Background:** Climate change, including global warming, is expected to cause poorer global health and a rise in the number of environmental refugees. As neurological disorders account for a major share of worldwide morbidity and mortality, climate change and global warming are also destined to alter neurological practice; however, to what extent and by which mechanisms is unknown. We aimed to collect the available information on the effects of ambient temperatures and human migration on the epidemiological and clinical manifestations of major neurological disorders.

**Methods:** We searched PubMed and Scopus from January 1, 2000 to November 30, 2020 for human studies published in English addressing the influence of ambient temperatures and human migration on Alzheimer’s and non-Alzheimer’s dementia, epilepsy, headache and migraine, multiple sclerosis, Parkinson’s disease, stroke, and tick-borne encephalitis (as a model disease for neuroinfections). The protocol was pre-registered at PROSPERO (2020 CRD42020147543).

**Results:** 101 studies met inclusion criteria, but we were unable to identify a single study addressing how global warming and human migration will change neurological practice. Still, extracted data suggested multiple ways by which these aspects might alter neurological morbidity and mortality in the future.

**Conclusion:** Significant heterogeneity exists across studies with respect to methodology, outcome measures, control of confounders and study design, but there is enough evidence to suggest climate change will affect the neurological practice of all major neurological disorders. Adequately designed studies to address this issue are urgently needed, which will require concerted efforts from the neurological community.

## INTRODUCTION

The United Nations have identified climate change, which includes global warming, as the “defining issue of our time”^1^. As stated in the Paris Climate Agreement, the increase of global temperature must be contained within 2, ideally 1.5, degrees Celsius above pre-industrial levels^2^ to avoid major negative consequences, including poorer global health and a rise of environmental refugees (i.e. displaced people from increasingly inhabitable regions of the world). Thus, climate change and global warming will likely worsen illnesses worldwide^3^ and may become the major drivers of human migration^4^.

According to the Global Burden of Disease Study, neurological disorders are the foremost cause of disability adjusted life years (DALYs), accounting for 10.2 percent of global DALYs, and neurological diseases are the second-leading cause of deaths, representing 16.8 percent of global deaths^5^. It is therefore reasonable to assume that climate change and global warming will also have a major impact on clinical neurological practice.

To start addressing this issue, one must first identify the areas of neurological practice that will likely be subject to alterations related to global warming. It is necessary to investigate in which ways a rise of ambient temperatures might affect the frequency, semiology, and outcome of major neurological disorders. Furthermore, the prevalence and incidence of neurological disorders in refugees and other human migrant populations might serve as a proxy for what could be expected in future environmental refugees.

In this review, our main objective was to identify how ambient temperatures influence the epidemiology and clinical aspects of major groups of neurological disorders. The second objective was to investigate the prevalence and incidence of neurological disorders in human refugee and migrant populations.

## METHODS

Using the PICO approach^6^, we phrased the following research questions:

1. In people with major neurological disorders, including Alzheimer’s and non-Alzheimer’s dementia, epilepsy, headache and migraine, multiple sclerosis, Parkinson’s disease, stroke, and tick-borne encephalitis (as a proxy for neuroinfectious diseases) (Population), how does an increase in ambient temperatures (Intervention), as compared to normal ambient temperatures (Comparison), affect the frequency, symptomatology, and mortality of these disorders (Outcome)?
2. In human refugee and migrant populations (Population), how does the fact of being displaced (Intervention), as compared to populations living in their home country (Comparison), affect the prevalence and incidence of neurological disorders (Outcome)?

For each PICO question a systematic review of the literature was performed using a predefined search. The full search strategy (including MeSH headings for searches in PubMed) is available from the ***Supplemental file 1***. The review was pre-registered at PROSPERO (2020 CRD42020147543).

### Types of studies

We evaluated all cross-sectional or longitudinal, retrospective or prospective, observational clinical and research studies on major neurological diseases with epidemiological and clinical data associated with ambient temperatures, as well as epidemiological and clinical data associated with human and refugee populations. We excluded interviews, editorials, opinions, and news articles.

### Electronic search strategy

We included human only studies published in English and listed in PubMed and Scopus from January 1^st^, 2000 to November 30, 2020. Non-English literature was included if an English abstract and a reliable translation of the manuscript into English were available. The literature search was supervised by an information specialist from the University of Copenhagen’s library service, and the search strategy was developed in accordance with the above PICO questions. For search examples and filters, the reader is referred to the ***Supplemental file 1***. The references of relevant articles were manually searched to identify additional articles. Further, papers were cross-referenced using the ‘cited by’ function in PubMed. When necessary, personal communication with authors was attempted by email or phone to obtain additional data.

### Data collection

After reviewing titles and abstracts, relevant studies were assessed on a full text basis. Data was extracted by the 1^st^, 2^nd^ and 3^rd^ authors and cross-checked by the senior author; any uncertainties concerning data extraction and interpretation were resolved by consensus.

### Statistical analyses

Owing to the high heterogeneity of the data including different definitions of temperature thresholds, quantitative statistical analysis was not performed.

## RESULTS

The primary search yielded 4,992 titles. After screening and removal of duplicates, 101 studies met the inclusion criteria for the final review (**Figure 1**). **Table 1** provides an overview of studies (n=85) regarding ambient temperatures and major neurological disorders. The effects of migration on prevalence and incidence of neurological disorders (n=16 studies) are outlined in **Tables 2** and **3**. Extracted raw data from the literature search are available on request from the corresponding author.

**TABLE 1.** Characteristics of studies included in this review, investigating the association between ambient temperature and major neurological disorders. Temperatures are given as maximum temperatures or increase in mean temperature. Only the effects of high temperatures on epidemiology, symptoms and mortality are listed

| **TABLE 1** |  | Characteristics of studies included in this review, investigating the association between ambient temperature and major neurological disorders. Temperatures are given as maximum temperatures or increase in mean temperature. Only the effects of high temperatures on epidemiology, symptoms and mortality are listed |  |  |  |  |  |  |
| --- | --- | --- | --- | --- | --- | --- | --- | --- |
| Diagnosis | Number of studies (%) | Study design | Site of study | Total case population | Temperature (maximal) | Effect on epidemiology (Increase in incidence/prevalence/hospitalization) | Worsening of symptoms | Mortality (increase) |
| Cerebrovascular disorders | 61 (70.1%) | Retrospective, observational | AU, CA, CN, DE, DK, ES, GB, IN, IL, IT, JP, KR, PR, QA, RU, SE, TU, TW, US | 5,869,284 | 12.9°C in 1 study<br>20-27°C in 8 studies<br>>30°C in 17 studies<br>N/A in 2 studies | 17 studies | N/A | 16 studies |
| Alzheimer and non-Alzheimer dementia | 10 (11.5%) | Retrospective, observational | AU, CN, ES, IT, KR, UK, US, VN | 960,675 | 18°C in 1 study<br>>24.5°C in 1 study<br>>30°C in 8 studies | 6 studies | 1 study | 3 studies |
| Multiple sclerosis | 6 (6.9%) | 5 studies: Retrospective, observational<br>1 study: Retro- and prospective, observational | AU, DE, FR, JP, US | 5,305 | >20°C | N/A | 4 studies | N/A |
| Headache and Migraine | 2 (2.3%) | Retrospective, observational | FR, US | 7,156 | Temperature increase > 5°C | 1 study | 1 study | N/A |
| Parkinson's disease | 3 (3.4%) | Retrospective, observational | ES, FR, US | 204,656 | >31.7°C in 3 studies | N/A | 2 study | 1 study |
| Epilepsy | 1 (1.1%) | Retrospective, observational | DE | 604 | >20°C | N/A | Decreased | N/A |
| Neuroinfectious disorders (TBE) | 4 (4.6%) | Retrospective, observational | CZ, SE, SI | N/A | N/A | 4 | N/A | N/A |
| Abbreviations: TBE: Tick-borne encephalitis; Temp: temperature N/A: Not applicable or available;<br>AU=Australia, CA=Canada, CN=China, CZ=Czech Republic, DE=Germany, DK=Denmark, ES=Spain, FR=France, GB=United Kingdom, IN=India, IS=Israel, IT=Italy, JP=Japan, KR=South Korea, PR=Puerto Rico, QA=Qatar, RU=Russia, SE=Sweden, SI=Slovenia, TR=Turkey, TW=Taiwan, US=United States, VN=Vietnam |  |  |  |  |  |  |  |  |

**TABLE 2.** Studies (n=8) investigating the differences in neurological disorders between migrants and populations from countries of origin

| TABLE 2 | Studies (n=8) investigating the differences in neurological disorders between migrants and populations from countries of origin |  |  |  |  |  |  |
| --- | --- | --- | --- | --- | --- | --- | --- |
| Article | Site | Study Design | Recruitment | Population (Participants, Sex, Age) | Country of origin → arrival | Effect on epidemiology (Increase in incidence/prevalence/hospitalization) | Mortality |
| **Cerebrovascular Disease** |  |  |  |  |  |  |  |
| <sup>e96</sup> | CN, HK, NAm, SG, TW, WEu, | Observational | Prospective | 680 CN, 69.6%M, 65.7±9y;<br>1,648 HK/SG/TW, 68%M, 65.5±9.8y;<br>169 WEu, 71%M, 67.3±9.1y;<br>441 NAm, 63.3%M, 69.8±10.4y | CN → HK/NAm/SG/TW/WEu | Decreased | No Effect |
| <sup>e95</sup> | DE, GB, GH, NL | Observational | Retrospective | 206 GH, 30.1%M, 52.86±9.9y;<br>444 DE/GB/NL, 50%M, 52.2±8.8y | GH → GB, DE, NL | Decreased | N/A |
| <sup>e97</sup> | BB, GB | Observational | Retrospective | 665 BB, 42.4%M, 71.2±14.9<br>271 GB, 66.1±13.7y | BB → GB | Increased | Increased |
| **Multiple Sclerosis** |  |  |  |  |  |  |  |
| <sup>e91</sup> | CA, IR | Observational | Retrospective | Onset of MS *before* migration:<br>29 IR, 20.1%M, 24 (19-31)y<br>Onset of MS *after* migration<br>48 IR, 33.3%M, 34 (25-40)y | IR → CA | Increased | N/A |
| <sup>e92</sup> | AU, GB, IE | Observational | Retrospective | 331 GB/IE, 208/331 with age 20-49y in 1981 | GB, IE → AU | Decreased | N/A |
| <sup>e93</sup> | Caribbean islands, MQ | Observational | Prospective | 53 Afro Caribbean, 13.2%M,<br>40.7±12.1y<br>59 MQ, 18.6%M, 43.2±10.4y | Caribbean → MQ | Decreased | N/A |
| <sup>e94</sup> | BR, IT, ES, PT | Observational | Retrospective | 652 BR/ES/IT/PT, 28.4%M, 42y | ES, IT, PT → BR | No Effect | N/A |
| **Dementia** |  |  |  |  |  |  |  |
| <sup>e90</sup> | BR, JP | Observational | Prospective | 157 JP, 44.6%M, 70-100y | JP → BR | Increased | Increased |
| Abbreviations: N/A: Not applicable or available; M=males; MS=multiple sclerosis; y=years;<br>AU=Australia, BB=Barbados, BR=Brazil, CN=China, CA=Canada, DE=Germany, ES=Spain, GB=United Kingdom, GH=Ghana, HK=Hong Kong, IE=Ireland, IR=Iran, IT=Italy, JP=Japan, MQ=Martinique NAm=North America, NL=Netherlands, PT=Portugal, SG=Singapore, TW=Taiwan, WEu=Western Europe |  |  |  |  |  |  |  |

**Figure 1.**
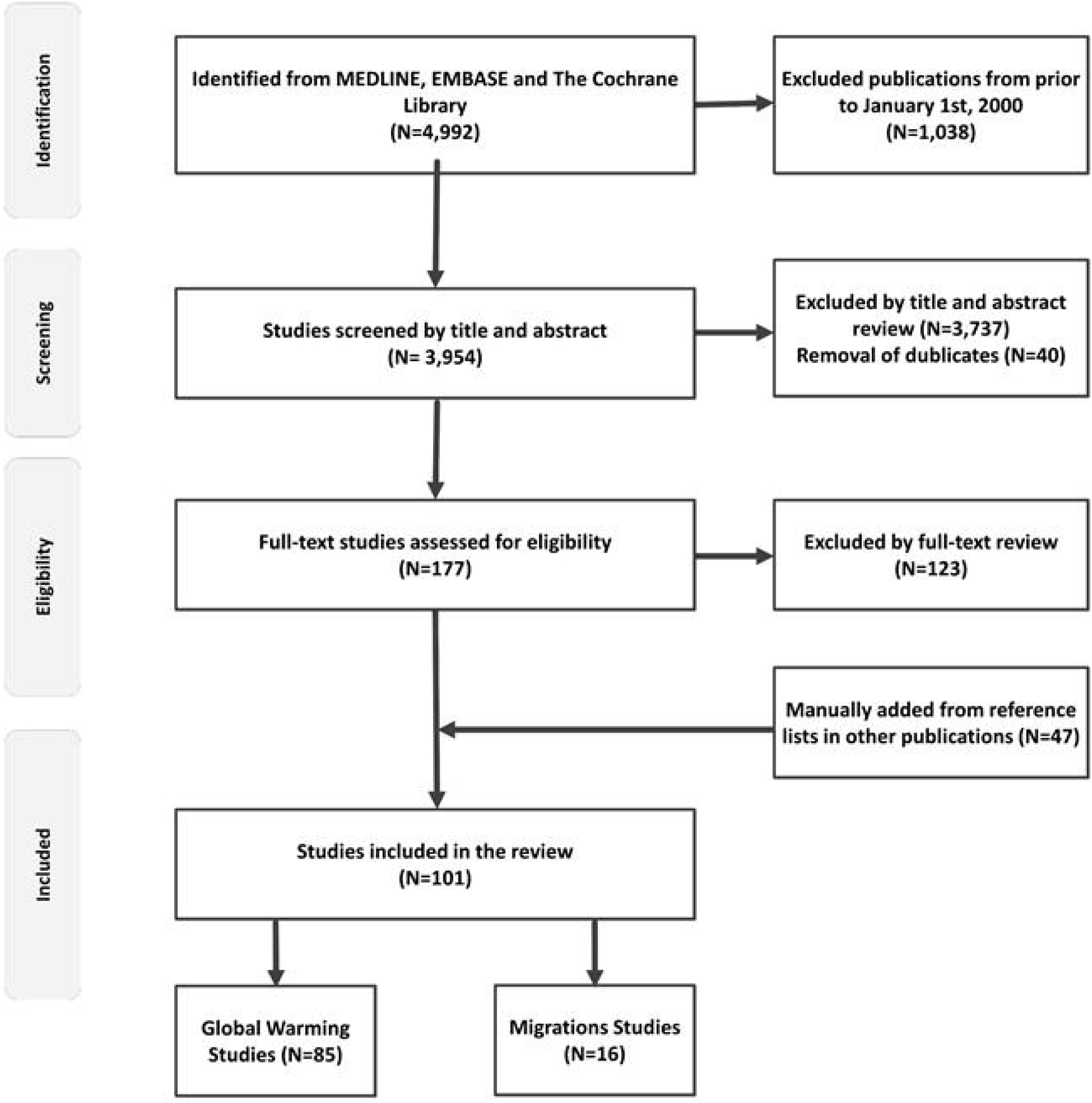
Flow chart of literature search

### Ambient temperature and major neurological disorders

#### Alzheimer’s and non-Alzheimer’s dementia

Ten studies investigated the effects of high ambient temperatures on patients with Alzheimer’s and non-Alzheimer’s dementia^7-16^. The studies were from 7 countries (AU, CN, ES, IT, KR, UK, US, and VN) with a total case population of N = 960,675. One study did not specify the number of cases^8^.

Maximum temperatures reported were >30°C in 8 studies^7,8,10-13,15-17^ and 18°C, respectively, >24.5°C in two other studies^9,14^. In 8 of the 10 studies, high ambient temperatures were associated with worsening of symptoms including agitation, as well as an increase in hospitalizations and/or mortality^7,8,10,12-17^. Five studies suggested that hot temperature were attributable to worsening of symptoms and increased risk of admission^7,12,14-16^, while Hansen et al. found a significantly increased rate of both hospital admissions and mortality in 94,447 patients with dementia during periods of heat waves^8^. Zanobetti et al, Xu et al. and Conti et al. also found significantly increased mortality during hot months^10,13,17^. In the remaining two studies, no associations were found between high ambient temperatures and hospital admissions or death in people with dementia ^9,11^. Overall, increased temperatures were associated with increased rates of hospitalization and mortality in patients with dementia in 8 of the 10 studies.

#### Epilepsy

Only one epilepsy study met inclusion criteria, a case-crossover study with 604 patients^18^. The results indicated a 46% lower risk of admissions for epileptic seizures one day after exposure to temperatures above 20°C. Further, an inverse association was found between epileptic seizures and low atmospheric pressure and high humidity^18^.

#### Headache and migraine

Two studies from 2 countries (US and FR) including N = 7,156 cases reported on the effects of high temperatures on headaches, including 33% migraine ^19,20^. The largest study by Mukamal et al. analyzed data from 7,054 patients seen in an emergency department between 2000-2007^19^. In this study, higher ambient temperatures in the 24 hours preceding hospital presentation increased the risk of acute headache requiring emergency evaluation with 7.5% for each 5°C increment in temperature. Neut et al. interviewed 102 children and adolescents with migraine and/or their parents about triggering factors precipitating migraine attacks. Seventy % stated warm climate could trigger their migraine, and 24% reported warm climate was often or very often a trigger factor for migraine attacks^20^.

#### Multiple sclerosis

Six studies from 5 countries (US, FR, DE, AU, JP) with a total N = 5,305 addressed the effects of high temperatures on patients with multiple sclerosis ^21-26^. Four studies found high temperatures to be associated with worsening of symptoms in these patients ^21,22,24,26^. Stellmann et al. included 1,254 patients and showed that deficits measured by a walking test slightly worsened when temperatures increased to 20°C^26^. In an anonymized survey by Simmons et al., 70% of multiple sclerosis participants reported worsening of their symptoms with high temperatures^22^. Poor cognitive performance significantly correlated with warmer outdoor temperatures in another group of 40 multiple sclerosis patients^24^. Furthermore, the latter study also found a significant correlation between poor cognitive performance and high temperatures over a period of 6 months, suggesting a negative impact of warm temperatures on cognitive function in multiple sclerosis patients when analyzed cross-sectionally and longitudinally. Higher frequencies of multiple sclerosis attacks were recorded during the warmest months in a small sample of 34 multiple sclerosis patients^21^. The remaining two studies did not show any effects of high temperatures on multiple sclerosis symptomatology ^23,25^. Thus, 4 of 6 studies reported worsening of symptoms in multiple sclerosis patients during warm periods.

#### Parkinson’s disease

Three studies from 3 countries (FR, ES and US) with a total N = 204,656 evaluated the effects of high temperatures on Parkinson’s disease ^10,27,28^. Results from one study indicated a correlation between high ambient temperature (>34°C) and an increase risk of excess morbidity and mortality in Parkinson patients^28^. In a small French study with 36 Parkinson’s disease patients, there was a trend towards more frequent symptoms of autonomic failure during heat wave periods^27^. In contrast, another study investigated the effects of extreme hot days (maximum temperature of 31,7°C) on mortality in 201,333 patients with a diagnosis of Parkinson’s disease but found no association between mortality and ambient temperatures^10^.

#### Stroke, overview

In total, 59 papers dealing with the effects of ambient temperatures on cerebrovascular morbidity and mortality were included ^10,29-60,e61-67^. The studies were from 18 countries (AU, CA, CN, DE, DK, ES, GB, IN, IL, IT, JP, KR, PR, QA, RU, SE, TU, TW, and US) with a total case population of N = 5,869,284. Ischemic stroke was reported in 1,812,457, hemorrhagic stroke in 446,407, and SAH in 7,160 cases. In the remaining 3,603,260 cases, stroke subtypes were not specified.

#### Stroke, hospitalizations

Associations between stroke-related hospitalization and ambient temperatures were reported in 38 studies. Thirteen studies reported increasing risks for hospitalizations for stroke with higher ambient temperatures ^29, 36, 44,46,50,56,e65-71^. Maximum temperatures were 12.9°C in one study^29^, 20-27°C in 5 studies^36,44,e65,69,70^, and >30°C in 5 studies^47,50,56,e66,71^. Temperatures were unspecified in two studies^e67, 68^. Chen et al. found excess hospitalizations due to ischemic stroke with high temperatures, while the risk for admissions because of hemorrhagic stroke was increased during both high and low temperatures^49^. Green et al. and Basu et al. also found increased risks of hospitalization due to ischemic stroke with high temperatures (25.3 and 30.1°C, respectively), but a decreased risk for hemorrhagic stroke^37, 42^. Bai et al. reported an increase in risk of stroke admissions both with high and low ambient temperatures^e62^. In 10 studies, lower ambient temperatures (−16.94 to 17 °C) were associated with increased risk of stroke-related hospitalizations^53,57,e64,72-78^. In the study of Wang et al., there was an inverse correlation between temperatures and ischemic stroke admissions, leading to an increase of admissions during cold spells and a decrease during heat waves^48^. Lin et al. reported lower risks of stroke admissions associated with high ambient temperatures (31.7°C), even though the results were not statistically significant^35^. Cevik et al. only found association between lower temperature and increased risk of SAH^e79^, and like the remaining eight studies no associations between ambient temperatures and stroke-related hospitalizations was found^30,34,41,43,47,59,e80,81^. Overall, most studies (17 of 24) reported higher risks of stroke-related hospitalizations during periods of high ambient temperatures.

#### Stroke, mortality

21 studies reported on the association between ambient temperatures and cerebrovascular mortality^10,31-33,38-40,45,51,52,54,55,58,e60,61,63,77,82-85^. In 11^10,31,32,38,39,45,55,e61,82-84^ of the 21 studies, increased risks of cerebrovascular death was associated with high ambient temperatures, including very high temperatures (>30°C)^10,31,32,38,39,55,e61,83,84^. Five studies reported an increase in cerebrovascular mortality with both low and high temperatures^51,52,58,60,e63^, while 2 studies found no effect^33,40^. Zhang et al., Myint et al. and Yang et al. were the only to report significantly higher risks of cerebrovascular death in periods with cold, but not warm, temperatures^54,e77,85^. Overall, most studies (16 of the 21 studies) found a higher risk of cerebrovascular mortality with increasing ambient temperatures.

#### Tick-borne encephalitis

The effects of high ambient temperatures on tick-borne encephalitis were assessed in 4 studies from 3 countries (SE, CZ, SI)^e86-89^. Lindgren et al. linked an increase in the incidence of tick-borne encephalitis in Sweden, starting in the mid-80s, to climate change with increasingly milder winters and earlier springs^e87^. Lukan et al. and Zeman et al. analyzed 1,786, respectively, 8,700 cases of tick-borne encephalitis in the period of 1961-70 to 2004^e88, 89^. During 1980 to 2004 they found an increase in tick-borne encephalitis foci in areas of increasing altitudes which corresponded to gradual rises in annual temperatures, indicating an effect of climate change and warmer temperatures on local tick-borne encephalitis incidence. Overall, the studies indicated a rise in the total incidence of tick-borne encephalitis with milder temperatures in Sweden and increasing temperatures in higher altitude areas in the Czech Republic and Slovenia.

### Neurological disorders in human migrant and refugee populations

Results from studies comparing migrants with populations from their country of origin are listed below and in **Table 2**.

#### Alzheimer’s and non-Alzheimer’s dementia

One observational study reported a higher prevalence of dementia in a group of elderly Japanese people (>70 years) who had migrated from Okinawa, Japan, to Brazil, as opposed to people who had stayed in Japan^e90^.

#### Multiple Sclerosis

Four observational studies investigated the effects of migration on the frequency^e91,92^ and morbidity^e93,94^ of multiple sclerosis. In the study by Guimond et al., multiple sclerosis prevalence was higher in migrants who came from Iran to British Columbia^91^, whereas in the study by Hammond et al. the incidence was higher in the population from the home country (UK and Ireland) compared to the incidence in a migrant population settling in Australia^e92^. Further, Merle et al. investigated visual impairment in multiple sclerosis patients from Martinique who had or had not migrated to metropolitan France for at least 1 year before the age of 15 years and found that symptoms were more frequent and severe in the non-migrant group^e93^. Comini-Frota et al. found no difference of multiple sclerosis morbidity between an Italian migrant population in Brazil and non-migrant Italians^e94^.

#### Stroke

Three observational studies compared data on cerebrovascular disease in migrant populations with data from non-migrant populations^e95-97^. Two studies reported a higher prevalence of stroke in non-migrant groups living in China^e96^ and Ghana^e95^, as opposed to migrant populations living in Hong Kong, Singapore, Taiwan, Western Europe, and North America. After adjusting for conventional cardiovascular risk factors, results were no longer significant in one study^e95^. In contrast, Wolfe et al. observed a higher incidence of stroke in migrants from Barbados moving to South London, including an increased incidence for specific stroke subtypes such as total anterior cerebral infarctions, posterior cerebral infarctions and subarachnoid hemorrhages^e97^.

Results from studies comparing migrants with populations from their country of arrival are summarized in **Table 3**.

**TABLE 3.** Studies (n=9) investigating the differences in neurological disorders between migrants and populations from countries of arrival

|  |  |  |  |  |  |  |  |
| --- | --- | --- | --- | --- | --- | --- | --- |
| Article | Site | Study Design | Recruitment | Population (Participants, Sex, Age) | Country of origin → arrival | Effect on epidemiology (Increase in incidence/prevalence/hospitalization) | Mortality |
| **Cerebrovascular Disease** |  |  |  |  |  |  |  |
| e120 | IT | Observational, Cross-sectional | Retrospective | 3,616.5 migrants (per 100.000), 50.9%M<br>3,281.1 IT (per 100.000), 50.1%M | Not specified → IT | Increased | N/A |
| e121 | GB | Observational | Prospective | 1,540 Caribbean, 25-54y | Caribbean → GB | N/A | Increased |
| e122 | US | Observational | Prospective | 746 Hispanic foreign-born, 41.8%M, 64.8y;<br>15,038 Hispanic and whites US-born, 44.1%M, 65.6y | Hispanic countries → US | Decreased | N/A |
| e123 | NO | Observational | Retrospective | 331 SA, 64.7%M;<br>229 FY, 70%M;<br>134 SSA, 82.8%M;<br>124,253 SEA, 0.06%M;<br>14 CAs, 42.9%M;<br>15 Cam, 0.4%M;<br>35-64y for all groups | 14 Regions → NO | SA: Increased<br>FY: Increased in men<br>SSA: Increased in men<br>SEA/CAs/CAM: Increased in women | N/A |
| e124 | FI, SE | Observational | Retrospective | 2,001 FI, 39%M, 64.7y;<br>1,574 SE, 36.6%M, 68.4y | FI → SE | N/A | Increased |
| e125 | CN, US | Observational, Cross-sectional | Retrospective | 11 CN, 38.1%M, 65.3±13.3<br>34 US, 43.1%M, 59.8±11.9y | CN → US | Decreased | N/A |
| **Multiple Sclerosis** |  |  |  |  |  |  |  |
| e126 | AU, GB, IE | Observational | Retrospective | 20 GB/IE, 30%M, 32.2y;<br>99 AU, 37.4%M, 32.6y | GB, IE → AU | No Effect | N/A |
| e94 | BR, IT, ES, PT | Observational | Retrospective | 652 BR/ES/IT/PT, 28.4%M, 42y | ES, IT, PT → BR | Increased in IT | N/A |
| **Headache and Migraine** |  |  |  |  |  |  |  |
| e127 | MX, US | Observational, Cross-sectional | Retrospective | 56 Mexican Americans, 23.2%M, >45y;<br>116 White Americans, 31%M, >45y;<br>41 Afro-Americans, 19.5%M, >45y | MX → US | No Effect | N/A |
| Abbreviations: Abbreviations: N/A: Not applicable or available; M=males; MS=multiple sclerosis; y=years;<br>AU=Australia, BR=Brazil, CAM=Central America, CAs=Central Asia, CN=China, ES=Spain, FI=Finland, FY=Former Yugoslavia GB=United Kingdom, IE=Ireland, IT=Italy, MX=Mexico, NO=Norway, PT=Portugal, SA=South Asia, SE=Sverige, SEA=Southeast Asia, SSA=Sub-Saharan Africa, US=United States |  |  |  |  |  |  |  |

## DISCUSSION

Although the United Nations have declared climate change the greatest challenge of the 21st century and although neurological disorders comprise the greatest share of DALYs globally, we were unable to identify a single adequately designed study addressing how climate change and its consequences will alter neurological practice in the future. This is concerning given that the evidence that we did find indeed suggests many ways by which the effects of global warming and human migration on neurological practice may unfold.

### Ambient temperatures and neurological disorders

Most stroke studies showed a relationship between increasing ambient temperatures and higher rates of hospitalization and mortality. Heat exposure may be associated with hemoconcentration and hyperviscosity, impaired endothelial function and hemodynamic disturbances, including cardiac arrhythmia, thereby increasing risks for both ischemic and hemorrhagic stroke^e98^.

Likewise, warmer temperatures were reported to worsen cognitive symptoms and to increase mortality in dementia patients. The pathogenesis is unclear, but impaired physiological adjustment to rising temperatures because of dysautonomia are known from patients with fronto-temporal dementia and Alzheimer’s disease and may cause dehydration, cardiorespiratory distress and susceptibility to drug side effects^e99^. Autonomic failure aggravated by high temperatures is also a plausible complication in Parkinson’s disease that may lead to orthostatic hypotension and ensuing risk for trauma from syncope and falls^27^.

With multiple sclerosis, most studies revealed a decline in motor and cognitive functions following a raise in ambient temperatures. While this is consistent with Uhthoff’s phenomenon, i.e. the well-known reversible aggravation of multiple sclerosis symptoms caused by the blocking or slowing of nerve conduction with heat, no studies have specifically addressed the importance of Uhthoff’s phenomenon relative to attack rates, inflammatory mechanisms and secondary neurodegeneration with rising temperatures. Furthermore, hot temperatures are trigger factors for headache and migraine^19,20^. Heat is known to cause vasodilation which may contribute to vascular migraine^e100, 101^, and water deprivation can provoke secondary headaches including migraine^e102^.

As to epilepsy, there are well-established relationships with temperature and pediatric febrile seizures, seizures in conjunction with heat strokes^e103^ and Dravet syndrome-related seizures associated with hot water; and increased body temperature may trigger hippocampal neuronal activity in mesial temporal lobe epilepsy^e104^. In addition, using tick-borne encephalitis as a model disease for neuroinfections, we found evidence that vector-borne diseases are prone to spread from endemic areas to currently non-endemic regions with increasing humidity and rising temperatures.

### Human migration and neurological disorders

The impact of migration on the prevalence, incidence and severity of major neurological disorders is substantial but not uniform. Social, economic and cultural characteristics of both the countries of origin and arrival influence results.

The higher prevalence of stroke in populations from mainland China compared to Western countries^e96^ may be due to higher dietary salt intake^e105^; poorly controlled hypertension^e106,107^ and less accessible healthcare^e96,108^. Furthermore, the higher stroke prevalence in Black Caribbean’s from South London might rely on socioeconomic factors and lifestyle changes that could unmask genetic susceptibilities^e97,109^, as Black Caribbean immigrants are more prone to hypertension and diabetes^e110-113^.

Similarly, environmental and genetic factors impact the prevalence and morbidity of multiple sclerosis in first-generation immigrants. With migration to higher risk countries, prevalence increases in the migrating population^e91^, whereas with migration from high to low risk countries, prevalence decreases^e92^. Genetic factors affect multiple sclerosis rates in the long term, in later generations, notably when massive immigration occurred^e94,114^.

Also, it must be borne in mind that the psychological trauma of being displaced, lack of employment, low socioeconomic status, poor housing conditions, social isolation and ethnic discrimination may have limited direct impact on neurological disease; but all these factors can worsen mood disorders and other psychiatric conditions and may prevent access to health care, which may have secondary effects on neurological morbidity and mortality^e115-117^.

### Current limitations and future directions

Heterogeneity related to study design, exposures, outcome measures, effect modifiers and data presentation limit comparison of study results. In addition, most studies identified in this review were based on retrospective data prone to selection bias. Also, how and where (e.g. outdoor versus indoor) ambient temperatures were measured was not standardized, and the range of temperatures investigated were wide. Studies did not account for factors that may influence temperatures such as humidity, air pollution and geography. In addition, studies comparing migrants to non-migrants were few and did not distinguish between first- and second-generation immigrants; and comparisons were made between migrant populations and either their countries of arrival or departure, but not both.

Importantly, the effect sizes related to global warming and human migration, i.e. exactly how much these factors will influence the morbidity and mortality of neurological disorders, are entirely unknown, as is the influence of other factors associated with climate change such as loss of biodiversity, rising sea levels and drought (**Figure 2**).

**Figure 2.**
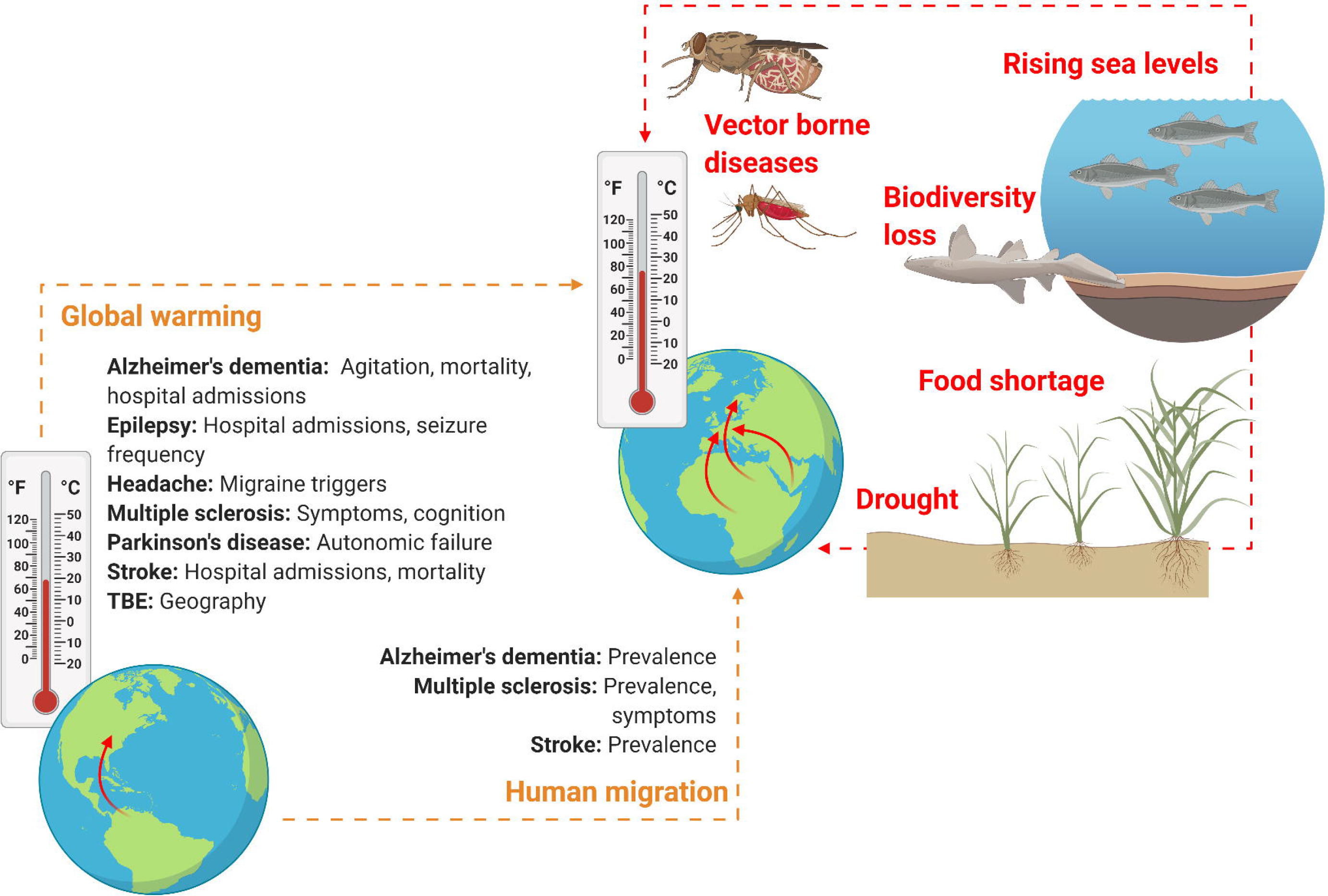
Schematic overview on how climate change might soon impact neurological practice. Global warming and human migration were covered in this Review (left). Although we identified no studies addressing precisely how and to what extent rising environmental temperatures may affect neurological disorders and only few studies that investigated neurological disorders in human migrant populations, it seems reasonable to assume that both global warming and climate refugees will alter clinical practice of various neurological disorders owing to alterations in prevalence, incidence, mortality, morbidity and disease semiology. However, global warming and human migration are only two aspects of climate change. Other factors (right) that may change neurological practice directly or indirectly and that were not addressed in this Review include drought, rising sea levels and loss of biodiversity (here, a dead nurse shark), which all might lead to altered neurological practice owing to e.g. food shortage, water insecurity and displacement of communities, as well as an increase in vector-borne diseases (here, a Tsetse fly and an Anopheles mosquito which are the vectors for African trypanosomiasis and cerebral malaria, respectively). *Figure created with Biorender.com*

Taking the lack of valid data into account, it is not surprising that, so far, predictions of the future epidemiology of neurological disorders are typically based on population growth and life expectancy but completely ignore the impact of climate change and its consequences^e118,119^.

In conclusion, although results were inconsistent due to the heterogeneity of data, our review suggests that climate change will soon change neurological practice because it affects morbidity and mortality of all major neurological disorders. Adequately designed studies to start addressing this issue are urgently needed, which will require coordinated efforts from the entire neurological community.

## Data Availability

Not applicable

## References

1. Climate Change [online]. Available at: https://www.un.org/en/sections/issues-depth/climate-change/. Accessed 17 November.

2. Paris agreement. United Nations2015.

3. Climate Change Impacts Human Health. United Nations Climate Change 2017.

4. Climate change and disaster displacement [online]. Accessed November 17.

5. Feigin VL, Abajobir AA, Abate KH, et al. Global, regional, and national burden of neurological disorders during 1990–2015: a systematic analysis for the Global Burden of Disease Study 2015. The Lancet Neurology 2017;16:877–897.

6. Schardt C, Adams MB, Owens T, Keitz S, Fontelo P. Utilization of the PICO framework to improve searching PubMed for clinical questions. BMC Med Inform Decis Mak 2007;7:16.

7. Cornali C, Franzoni S, Riello R, Ghianda D, Frisoni GB, Trabucchi M. Effect of high climate temperature on the behavioral and psychological symptoms of dementia. J Am Med Dir Assoc 2004;5:161–166.

8. Hansen A, Bi P, Nitschke M, Ryan P, Pisaniello D, Tucker G. The Effect of Heat Waves on Mental Health in a Temperate Australian City. Environmental Health Perspectives 2008;116:1369–1375.

9. Page LA, Hajat S, Kovats RS, Howard LM. Temperature-related deaths in people with psychosis, dementia and substance misuse. Br J Psychiatry 2012;200:485–490.

10. Zanobetti A, O’Neill MS, Gronlund CJ, Schwartz JD. Susceptibility to mortality in weather extremes: effect modification by personal and small-area characteristics. Epidemiology 2013;24:809–819.

11. Trang PM, Rocklöv J, Giang KB, Kullgren G, Nilsson M. Heatwaves and Hospital Admissions for Mental Disorders in Northern Vietnam. PLoS One 2016;11:e0155609.

12. Culqui DR, Linares C, Ortiz C, Carmona R, Díaz J. Association between environmental factors and emergency hospital admissions due to Alzheimer’s disease in Madrid. Sci Total Environ 2017;592:451–457.

13. Conti S, Masocco M, Meli P, et al. General and specific mortality among the elderly during the 2003 heat wave in Genoa (Italy). Environ Res 2007;103:267–274.

14. Lee S, Lee H, Myung W, Kim EJ, Kim H. Mental disease-related emergency admissions attributable to hot temperatures. Sci Total Environ 2018;616-617:688–694.

15. Linares C, Culqui D, Carmona R, Ortiz C, Díaz J. Short-term association between environmental factors and hospital admissions due to dementia in Madrid. Environ Res 2017;152:214–220.

16. Zhang Y, Nitschke M, Krackowizer A, et al. Risk factors of direct heat-related hospital admissions during the 2009 heatwave in Adelaide, Australia: a matched case-control study. BMJ Open 2016;6:e010666.

17. Xu Z, Tong S, Cheng J, et al. Heatwaves, hospitalizations for Alzheimer’s disease, and postdischarge deaths: A population-based cohort study. Environ Res 2019;178:108714.

18. Rakers F, Walther M, Schiffner R, et al. Weather as a risk factor for epileptic seizures: A case-crossover study. Epilepsia 2017;58:1287–1295.

19. Mukamal KJ, Wellenius GA, Suh HH, Mittleman MA. Weather and air pollution as triggers of severe headaches. Neurology 2009;72:922–927.

20. Neut D, Fily A, Cuvellier JC, Vallée L. The prevalence of triggers in paediatric migraine: a questionnaire study in 102 children and adolescents. J Headache Pain 2012;13:61–65.

21. Ogawa G, Mochizuki H, Kanzaki M, Kaida K, Motoyoshi K, Kamakura K. Seasonal variation of multiple sclerosis exacerbations in Japan. Neurol Sci 2004;24:417–419.

22. Simmons RD, Ponsonby AL, van der Mei IA, Sheridan P. What affects your MS? Responses to an anonymous, Internet-based epidemiological survey. Mult Scler 2004;10:202–211.

23. Tataru N, Vidal C, Decavel P, Berger E, Rumbach L. Limited impact of the summer heat wave in France (2003) on hospital admissions and relapses for multiple sclerosis. Neuroepidemiology 2006;27:28–32.

24. Leavitt VM, Sumowski JF, Chiaravalloti N, Deluca J. Warmer outdoor temperature is associated with worse cognitive status in multiple sclerosis. Neurology 2012;78:964–968.

25. Roberg BL, Bruce JM. Reconsidering outdoor temperature and cognition in multiple sclerosis. Mult Scler 2016;22:694–697.

26. Stellmann JP, Young KL, Vettorazzi E, Pöttgen J, Heesen C. No relevant impact of ambient temperature on disability measurements in a large cohort of patients with multiple sclerosis. Eur J Neurol 2017;24:851–857.

27. Pathak A, Lapeyre-Mestre M, Montastruc JL, Senard JM. Heat-related morbidity in patients with orthostatic hypotension and primary autonomic failure. Mov Disord 2005;20:1213–1219.

28. Linares C, Martinez-Martin P, Rodríguez-Blázquez C, Forjaz MJ, Carmona R, Díaz J. Effect of heat waves on morbidity and mortality due to Parkinson’s disease in Madrid: A time-series analysis. Environ Int 2016;89-90:1–6.

29. Dawson J, Weir C, Wright F, et al. Associations between meteorological variables and acute stroke hospital admissions in the west of Scotland. Acta Neurol Scand 2008;117:85–89.

30. Feigin VL, Nikitin YP, Bots ML, Vinogradova TE, Grobbee DE. A population-based study of the associations of stroke occurrence with weather parameters in Siberia, Russia (1982–92). European Journal of Neurology 2000;7:171–178.

31. Stafoggia M, Forastiere F, Agostini D, et al. Factors affecting in-hospital heat-related mortality: a multi-city case-crossover analysis. J Epidemiol Community Health 2008;62:209–215.

32. Vaneckova P, Hart MA, Beggs PJ, de Dear RJ. Synoptic analysis of heat-related mortality in Sydney, Australia, 1993-2001. Int J Biometeorol 2008;52:439–451.

33. Basu R, Ostro BD. A Multicounty Analysis Identifying the Populations Vulnerable to Mortality Associated with High Ambient Temperature in California. American Journal of Epidemiology 2008;168:632–637.

34. Knowlton K, Rotkin-Ellman M, King G, et al. The 2006 California heat wave: impacts on hospitalizations and emergency department visits. Environ Health Perspect 2009;117:61–67.

35. Lin S, Luo M, Walker RJ, Liu X, Hwang SA, Chinery R. Extreme high temperatures and hospital admissions for respiratory and cardiovascular diseases. Epidemiology 2009;20:738–746.

36. Wang XY, Barnett AG, Hu W, Tong S. Temperature variation and emergency hospital admissions for stroke in Brisbane, Australia, 1996-2005. Int J Biometeorol 2009;53:535–541.

37. Green RS, Basu R, Malig B, Broadwin R, Kim JJ, Ostro B. The effect of temperature on hospital admissions in nine California counties. Int J Public Health 2010;55:113–121.

38. Huang W, Kan H, Kovats S. The impact of the 2003 heat wave on mortality in Shanghai, China. The Science of the total environment 2010;408:2418–2420.

39. Basagaña X, Sartini C, Barrera-Gómez J, et al. Heat waves and cause-specific mortality at all ages. Epidemiology 2011;22:765–772.

40. Wichmann J, Andersen ZJ, Ketzel M, Ellermann T, Loft S. Apparent temperature and cause-specific mortality in Copenhagen, Denmark: a case-crossover analysis. Int J Environ Res Public Health 2011;8:3712–3727.

41. Wichmann J, Andersen Z, Ketzel M, Ellermann T, Loft S. Apparent temperature and cause-specific emergency hospital admissions in Greater Copenhagen, Denmark. PLoS One 2011;6:e22904.

42. Basu R, Pearson D, Malig B, Broadwin R, Green R. The effect of high ambient temperature on emergency room visits. Epidemiology 2012;23:813–820.

43. Cowperthwaite MC, Burnett MG. An analysis of admissions from 155 United States hospitals to determine the influence of weather on stroke incidence. Journal of Clinical Neuroscience 2011;18:618–623.

44. Ha S, Talbott EO, Kan H, Prins CA, Xu X. The effects of heat stress and its effect modifiers on stroke hospitalizations in Allegheny County, Pennsylvania. Int Arch Occup Environ Health 2014;87:557–565.

45. Lim Y-H, Kim H, Hong Y-C. Variation in mortality of ischemic and hemorrhagic strokes in relation to high temperature. International Journal of Biometeorology 2013;57:145–153.

46. Shaposhnikov D, Revich B, Gurfinkel Y, Naumova E. The influence of meteorological and geomagnetic factors on acute myocardial infarction and brain stroke in Moscow, Russia. Int J Biometeorol 2014;58:799–808.

47. Vaneckova P, Bambrick H. Cause-specific hospital admissions on hot days in Sydney, Australia. PLoS One 2013;8:e55459.

48. Wang Q, Gao C, Wang H, Lang L, Yue T, Lin H. Ischemic stroke hospital admission associated with ambient temperature in Jinan, China. PLoS One 2013;8:e80381.

49. Chen J-h, Jiang H, Wu L, et al. Association of ischemic and hemorrhagic strokes hospital admission with extreme temperature in Nanchang, China—A case-crossover study. Journal of Clinical Neuroscience 2017;43:89–93.

50. Chen T, Sarnat SE, Grundstein AJ, Winquist A, Chang HH. Time-series Analysis of Heat Waves and Emergency Department Visits in Atlanta, 1993 to 2012. Environ Health Perspect 2017;125:057009.

51. Chen R, Wang C, Meng X, et al. Both low and high temperature may increase the risk of stroke mortality. Neurology 2013;81:1064–1070.

52. Chen R, Yin P, Wang L, et al. Association between ambient temperature and mortality risk and burden: time series study in 272 main Chinese cities. BMJ 2018;363:k4306.

53. Wang YC, Lin YK. Association between temperature and emergency room visits for cardiorespiratory diseases, metabolic syndrome-related diseases, and accidents in metropolitan Taipei. PLoS One 2014;9:e99599.

54. Zhang Y, Li S, Pan X, et al. The effects of ambient temperature on cerebrovascular mortality: an epidemiologic study in four climatic zones in China. Environ Health 2014;13:24.

55. Méndez-Lázaro P, Pérez-Cardona C, Rodríguez E, et al. Climate change, heat, and mortality in the tropical urban area of San Juan, Puerto Rico. International journal of biometeorology 2016;62.

56. Vodonos A, Novack V, Horev A, Salameh I, Lotan Y, Ifergane G. Do Gender and Season Modify the Triggering Effect of Ambient Temperature on Ischemic Stroke? Women’s health issues : official publication of the Jacobs Institute of Women’s Health 2016;27.

57. Guo P, Zheng M, Wang Y, et al. Effects of ambient temperature on stroke hospital admissions: Results from a time-series analysis of 104,432 strokes in Guangzhou, China. Sci Total Environ 2017;580:307–315.

58. Han J, Liu S, Zhang J, et al. The impact of temperature extremes on mortality: A time-series study in Jinan, China. BMJ Open 2017;7:e014741.

59. Ponjoan A, Blanch J, Alves-Cabratosa L, et al. Effects of extreme temperatures on cardiovascular emergency hospitalizations in a Mediterranean region: a self-controlled case series study. Environmental Health 2017;16:32.

60. Zhang Y, Li C, Feng R, et al. The Short-Term Effect of Ambient Temperature on Mortality in Wuhan, China: A Time-Series Study Using a Distributed Lag Non-Linear Model. Int J Environ Res Public Health 2016;13.

61. Zhou L, Chen K, Chen X, et al. Heat and mortality for ischemic and hemorrhagic stroke in 12 cities of Jiangsu Province, China. Science of The Total Environment 2017;601602:271–277.

62. Bai L, Li Q, Wang J, et al. Increased coronary heart disease and stroke hospitalisations from ambient temperatures in Ontario. Heart 2018;104:673–679.

63. Fu SH, Gasparrini A, Rodriguez PS, Jha P. Mortality attributable to hot and cold ambient temperatures in India: a nationally representative case-crossover study. PLOS Medicine 2018;15:e1002619.

64. Luo Y, Li H, Huang F, et al. The cold effect of ambient temperature on ischemic and hemorrhagic stroke hospital admissions: A large database study in Beijing, China between years 2013 and 2014—Utilizing a distributed lag non-linear analysis. Environmental Pollution 2018;232:90–96.

65. Sherbakov T, Malig B, Guirguis K, Gershunov A, Basu R. Ambient temperature and added heat wave effects on hospitalizations in California from 1999 to 2009. Environ Res 2018;160:83–90.

66. Bao J, Guo Y, Wang Q, et al. Effects of heat on first-ever strokes and the effect modification of atmospheric pressure: A time-series study in Shenzhen, China. Sci Total Environ 2019;654:1372–1378.

67. Shimomura R, Hosomi N, Tsunematsu M, et al. Warm Front Passage on the Previous Day Increased Ischemic Stroke Events. J Stroke Cerebrovasc Dis 2019;28:1873–1878.

68. Ostro B, Rauch S, Green R, Malig B, Basu R. The effects of temperature and use of air conditioning on hospitalizations. Am J Epidemiol 2010;172:1053–1061.

69. Tarnoki AD, Turker A, Tarnoki DL, et al. Relationship between weather conditions and admissions for ischemic stroke and subarachnoid hemorrhage. Croat Med J 2017;58:56–62.

70. Oudin A, Strömberg U, Jakobsson K, Stroh E, Björk J. Estimation of short-term effects of air pollution on stroke hospital admissions in southern Sweden. Neuroepidemiology 2010;34:131–142.

71. Salam A, Kamran S, Bibi R, et al. Meteorological Factors and Seasonal Stroke Rates: A Four-year Comprehensive Study. J Stroke Cerebrovasc Dis 2019;28:2324–2331.

72. Goggins WB, Woo J, Ho S, Chan EY, Chau PH. Weather, season, and daily stroke admissions in Hong Kong. Int J Biometeorol 2012;56:865–872.

73. Mostofsky E, Wilker EH, Schwartz J, et al. Short-term changes in ambient temperature and risk of ischemic stroke. Cerebrovasc Dis Extra 2014;4:9–18.

74. Hori A, Hashizume M, Tsuda Y, Tsukahara T, Nomiyama T. Effects of weather variability and air pollutants on emergency admissions for cardiovascular and cerebrovascular diseases. Int J Environ Health Res 2012;22:416–430.

75. Matsumoto M, Ishikawa S, Kajii E. Cumulative effects of weather on stroke incidence: a multi-community cohort study in Japan. J Epidemiol 2010;20:136–142.

76. Hong YC, Rha JH, Lee JT, Ha EH, Kwon HJ, Kim H. Ischemic stroke associated with decrease in temperature. Epidemiology 2003;14:473–478.

77. Myint PK, Vowler SL, Woodhouse PR, Redmayne O, Fulcher RA. Winter excess in hospital admissions, in-patient mortality and length of acute hospital stay in stroke: a hospital database study over six seasonal years in Norfolk, UK. Neuroepidemiology 2007;28:79–85.

78. Zheng Y, Wang X, Liu J, Zhao F, Zhang J, Feng H. A Community-Based Study of the Correlation of Hemorrhagic Stroke Occurrence with Meteorologic Factors. J Stroke Cerebrovasc Dis 2016;25:2323–2330.

79. Çevik Y, Doğan N, Daş M, Ahmedali A, Kul S, Bayram H. The association between weather conditions and stroke admissions in Turkey. Int J Biometeorol 2015;59:899–905.

80. Kyobutungi C, Grau A, Stieglbauer G, Becher H. Absolute temperature, temperature changes and stroke risk: a case-crossover study. Eur J Epidemiol 2005;20:693–698.

81. Bobb JF, Obermeyer Z, Wang Y, Dominici F. Cause-specific risk of hospital admission related to extreme heat in older adults. JAMA 2014;312:2659–2667.

82. Yang J, Zhou M, Li M, et al. Vulnerability to the impact of temperature variability on mortality in 31 major Chinese cities. Environ Pollut 2018;239:631–637.

83. Harlan SL, Chowell G, Yang S, et al. Heat-related deaths in hot cities: estimates of human tolerance to high temperature thresholds. Int J Environ Res Public Health 2014;11:3304–3326.

84. Qian Z, He Q, Lin HM, et al. High temperatures enhanced acute mortality effects of ambient particle pollution in the “oven” city of Wuhan, China. Environ Health Perspect 2008;116:1172–1178.

85. Yang J, Yin P, Zhou M, et al. The burden of stroke mortality attributable to cold and hot ambient temperatures: Epidemiological evidence from China. Environ Int 2016;92-93:232–238.

86. Danielová V, Kliegrová S, Daniel M, Benes C. Influence of climate warming on tickborne encephalitis expansion to higher altitudes over the last decade (1997-2006) in the Highland Region (Czech Republic). Cent Eur J Public Health 2008;16:4–11.

87. Lindgren E, Gustafson R. Tick-borne encephalitis in Sweden and climate change. Lancet 2001;358:16–18.

88. Lukan M, Bullova E, Petko B. Climate warming and tick-borne encephalitis, Slovakia. Emerg Infect Dis 2010;16:524–526.

89. Zeman P, Bene C. A tick-borne encephalitis ceiling in Central Europe has moved upwards during the last 30 years: possible impact of global warming? Int J Med Microbiol 2004;293 Suppl 37:48–54.

90. Yamada T, Kadekaru H, Matsumoto S, et al. Prevalence of dementia in the older Japanese-Brazilian population. Psychiatry and clinical neurosciences 2002;56:71–75.

91. Guimond C, Lee JD, Ramagopalan SV, et al. Multiple sclerosis in the Iranian immigrant population of BC, Canada: prevalence and risk factors. Mult Scler 2014;20:1182–1188.

92. Hammond SR, English DR, McLeod JG. The age-range of risk of developing multiple sclerosis: evidence from a migrant population in Australia. Brain 2000;123 (Pt 5):968–974.

93. Merle H, Smadja D, Merle S, et al. Visual phenotype of multiple sclerosis in the Afro-Caribbean population and the influence of migration to metropolitan France. Eur J Ophthalmol 2005;15:392–399.

94. Comini-Frota ER, Brum DG, Kaimen-Maciel DR, Fragoso YD, Barreira AA, Donadi EA. Frequency of reported European ancestry among multiple sclerosis patients from four cities in the southern and southeastern regions of Brazil. Clin Neurol Neurosurg 2013;115:1642–1646.

95. Hayfron-Benjamin C, van den Born BJ, Maitland-van der Zee AH, et al. Microvascular and macrovascular complications in type 2 diabetes Ghanaian residents in Ghana and Europe: The RODAM study. J Diabetes Complications 2019;33:572–578.

96. Chiu JF, Bell AD, Herman RJ, et al. Cardiovascular risk profiles and outcomes of Chinese living inside and outside China. Eur J Cardiovasc Prev Rehabil 2010;17:668–675.

97. Wolfe CD, Corbin DO, Smeeton NC, et al. Estimation of the risk of stroke in black populations in Barbados and South London. Stroke 2006;37:1986–1990.

98. Lavados PM, Olavarría VV, Hoffmeister L. Ambient Temperature and Stroke Risk: Evidence Supporting a Short-Term Effect at a Population Level From Acute Environmental Exposures. Stroke 2018;49:255–261.

99. Fletcher PD, Downey LE, Golden HL, et al. Pain and temperature processing in dementia: a clinical and neuroanatomical analysis. Brain 2015;138:3360–3372.

100. Goadsby PJ. Pathophysiology of migraine. Neurol Clin 2009;27:335–360.

101. Shevel E. The Extracranial Vascular Theory of Migraine-A Great Story Confirmed by the Facts. Headache 2011;51:409–417.

102. Blau JN. Water deprivation: a new migraine precipitant. Headache 2005;45:757–759.

103. Leon LR, Bouchama A. Heat stroke. Compr Physiol 2015;5:611–647.

104. Wieser HG. Mesial Temporal Lobe Epilepsy with Hippocampal Sclerosis. Epilepsia 2004;45:695-

105. Zhao L, Stamler J, Yan L, et al. Blood Pressure Differences Between Northern and Southern Chinese: Role of Dietary Factors The International Study on Macronutrients and Blood Pressure. Hypertension 2004;43:1332–1337.

106. Gong Z, Zhao D. Cardiovascular diseases and risk factors among Chinese immigrants. Intern Emerg Med 2016;11:307–318.

107. Huang X-B, Zhang Y, Wang T-D, et al. Prevalence, awareness, treatment, and control of hypertension in southwestern China. Scientific Reports 2019;9:19098.

108. Song S, Yuan B, Zhang L, et al. Increased Inequalities in Health Resource and Access to Health Care in Rural China. Int J Environ Res Public Health 2018;16.

109. Parlevliet JL, Uysal-Bozkir Ö, Goudsmit M, et al. Prevalence of mild cognitive impairment and dementia in older non-western immigrants in the Netherlands: a cross-sectional study. Int J Geriatr Psychiatry 2016;31:1040–1049.

110. Smeeton NC, Corbin DO, Hennis AJ, et al. Differences in risk factors between black Caribbean patients with stroke in Barbados and South london. Stroke 2009;40:640–643.

111. Charles A, van Oeffelen A, Norredam M, et al. Socioeconomic Inequalities in Stroke Incidence Among Migrant Groups Analysis of Nationwide Data. Stroke 2014;45.

112. Bidulescu A, Francis DK, Ferguson TS, et al. Disparities in hypertension among black Caribbean populations: a scoping review by the U.S. Caribbean Alliance for Health Disparities Research Group (USCAHDR). Int J Equity Health 2015;14:125.

113. Lane D, Beevers DG, Lip GYH. Ethnic differences in blood pressure and the prevalence of hypertension in England. Journal of Human Hypertension 2002;16:267–273.

114. Sloka JS, Pryse-Phillips WEM, Stefanelli M. Multiple Sclerosis in Newfoundland and Labrador - A Model for Disease Prevalence. Canadian Journal of Neurological Sciences / Journal Canadien des Sciences Neurologiques 2005;32:43–49.

115. Virupaksha HG, Kumar A, Nirmala BP. Migration and mental health: An interface. J Nat Sci Biol Med 2014;5:233–239.

116. Selten J-P, Termorshuizen F, van Sonsbeek M, Bogers J, Schmand B. Migration and dementia: a meta-analysis of epidemiological studies in Europe. Psychological Medicine 2020:1–8.

117. Selten JP, van der Ven E, Termorshuizen F. Migration and psychosis: a meta-analysis of incidence studies. Psychol Med 2020;50:303–313.

118. Dorsey ER, Constantinescu R, Thompson JP, et al. Projected number of people with Parkinson disease in the most populous nations, 2005 through 2030. Neurology 2007;68:384–386.

119. Wafa HA, Wolfe CDA, Emmett E, Roth GA, Johnson CO, Wang Y. Burden of Stroke in Europe: Thirty-Year Projections of Incidence, Prevalence, Deaths, and Disability-Adjusted Life Years. Stroke 2020;51:2418–2427.

120. de Waure C, Bruno S, Furia G, et al. Health inequalities: an analysis of hospitalizations with respect to migrant status, gender and geographical area. BMC International Health and Human Rights 2015;15:2.

121. Harding S. Mortality of migrants from the Caribbean to England and Wales: effect of duration of residence. International Journal of Epidemiology 2004;33:382–386.

122. Moon JR, Capistrant BD, Kawachi I, et al. Stroke incidence in older US Hispanics: is foreign birth protective? Stroke 2012;43:1224–1229.

123. Rabanal KS, Selmer RM, Igland J, Tell GS, Meyer HE. Ethnic inequalities in acute myocardial infarction and stroke rates in Norway 1994-2009: a nationwide cohort study (CVDNOR). BMC Public Health 2015;15:1073.

124. Albin B, Hjelm K, Elmståhl S. Comparison of stroke mortality in Finnish-born migrants living in Sweden 1970-1999 and in Swedish-born individuals. J Immigr Minor Health 2014;16:18–23.

125. Corlin L, Woodin M, Thanikachalam M, Lowe L, Brugge D. Evidence for the healthy immigrant effect in older Chinese immigrants: a cross-sectional study. BMC Public Health 2014;14:603.

126. Barnett MH, McLeod JG, Hammond SR, Kurtzke JF. Migration and multiple sclerosis in immigrants from United Kingdom and Ireland to Australia: a reassessment. III: risk of multiple sclerosis in UKI immigrants and Australian-born in Hobart, Tasmania. J Neurol 2016;263:792–798.

127. Molgaard CA, Rothrock J, Stang PE, Golbeck AL. Prevalence of migraine among Mexican Americans in San Diego, California: survey 1. Headache 2002;42:878–882.

