## Supplemental file for "Global Warming and Neurological Practice: Systematic Review"

**Supplemental File 1:**

***Electronic literature search***

We will search PubMed and Embase for existing English literature until July 31^st^, 2019. (The search will be updated shortly before submission of the planned manuscript in order to include also the newest references.) The literature search will be supervised by an information specialist from the Copenhagen University Library Service, and the search strategy will be developed in accordance with the clinical question.

For objective a), we will use the following search terms/MeSH terms: “headache”, OR “headache disorders”, “migraine”, “Alzheimer’s disease”, “dementia”, “stroke”, “cerebrovascular accident”, “heat stroke”, “epilepsy”, “central nervous system infections”, “Parkinson’s disease”, “multiple sclerosis”, AND “global warming”, “greenhouse effect” “heat wave”, “hot temperature”, “extreme heat”.

For objective b), we will use the following search terms/ MeSH terms: “refugees”, OR “transient and migrants”, “migrant”, “human migration”, “emigration and immigration”, “asylum seeker”, AND “headache”, OR “headache disorders”, “migraine”, “Alzheimer’s disease”, “dementia”, “stroke”, “cerebrovascular accident”, “heat stroke”, “epilepsy”, “central nervous system infections”, “Parkinson’s disease”, “multiple sclerosis”.

The references of relevant articles will be manually searched to identify additional articles. Further, papers will be cross-referenced using the ‘cited by’ function on PubMed. If necessary, personal communication with authors will be attempted via email or phone in order to obtain additional relevant data. The search strategies (including MeSH headings for searches in PubMed) will be saved and recorded in an appendix.

***Full electronic search strategy for Pubmed:***

(((((("global warming"[MeSH Terms] OR ("global"[All Fields] AND "warming"[All Fields]) OR "global warming"[All Fields]) OR ("greenhouse effect"[MeSH Terms] OR ("greenhouse"[All Fields] AND "effect"[All Fields]) OR "greenhouse effect"[All Fields])) OR heat wave[Text Word]) OR ("hot temperature"[MeSH Terms] OR ("hot"[All Fields] AND "temperature"[All Fields]) OR "hot temperature"[All Fields])) OR ("extreme heat"[MeSH Terms] OR ("extreme"[All Fields] AND "heat"[All Fields]) OR "extreme heat"[All Fields])) OR ((((("refugees"[MeSH Terms] OR "refugees"[All Fields]) OR ("transients and migrants"[MeSH Terms] OR ("transients"[All Fields] AND "migrants"[All Fields]) OR "transients and migrants"[All Fields])) OR ("human migration"[MeSH Terms] OR "human migration"[All Fields]) OR ("emigration and immigration"[MeSH Terms] OR ("emigration"[All Fields] AND "immigration"[All Fields]) OR "emigration and immigration"[All Fields])) OR ("refugees"[MeSH Terms] OR "refugees"[All Fields] OR ("asylum"[All Fields] AND "seeker"[All Fields]) OR "asylum seeker"[All Fields])) OR ("transients and migrants"[MeSH Terms] OR ("transients"[All Fields] AND "migrants"[All Fields]) OR "transients and migrants"[All Fields] OR "migrant"[All Fields]))) AND (((((((((((("headache"[MeSH Terms] OR "headache"[All Fields]) OR ("headache disorders"[MeSH Terms] OR ("headache"[All Fields] AND "disorders"[All Fields]) OR "headache disorders"[All Fields])) OR ("migraine disorders"[MeSH Terms] OR ("migraine"[All Fields] AND "disorders"[All Fields]) OR "migraine disorders"[All Fields] OR "migraine"[All Fields])) OR ("alzheimer disease"[MeSH Terms] OR ("alzheimer"[All Fields] AND "disease"[All Fields]) OR "alzheimer disease"[All Fields] OR ("alzheimer's"[All Fields] AND "disease"[All Fields]) OR "alzheimer's disease"[All Fields])) OR ("dementia"[MeSH Terms] OR "dementia"[All Fields])) OR ("stroke"[MeSH Terms] OR "stroke"[All Fields])) OR ("stroke"[MeSH Terms] OR "stroke"[All Fields] OR ("cerebrovascular"[All Fields] AND "accident"[All Fields]) OR "cerebrovascular accident"[All Fields])) OR ("heat stroke"[MeSH Terms] OR ("heat"[All Fields] AND "stroke"[All Fields]) OR "heat stroke"[All Fields])) OR ("epilepsy"[MeSH Terms] OR "epilepsy"[All Fields])) OR ("central nervous system infections"[MeSH Terms] OR ("central"[All Fields] AND "nervous"[All Fields] AND "system"[All Fields] AND "infections"[All Fields]) OR "central nervous system infections"[All Fields])) OR ("parkinson disease"[MeSH Terms] OR ("parkinson"[All Fields] AND "disease"[All Fields]) OR "parkinson disease"[All Fields] OR ("parkinson's"[All Fields] AND "disease"[All Fields]) OR "parkinson's disease"[All Fields])) OR ("multiple sclerosis"[MeSH Terms] OR ("multiple"[All Fields] AND "sclerosis"[All Fields]) OR "multiple sclerosis"[All Fields])) AND "humans"[MeSH Terms]
